# Acute upper airway disease in children with the omicron (B.1.1.529) variant of SARS-CoV-2: a report from the National COVID Cohort Collaborative (N3C)

**DOI:** 10.1101/2022.01.27.22269865

**Authors:** Blake Martin, Peter E. DeWitt, Seth Russell, L. Nelson Sanchez-Pinto, Melissa A. Haendel, Richard Moffitt, Tellen D. Bennett

## Abstract

**Background:** Reports of SARS-CoV-2 causing laryngotracheobronchitis (commonly known as croup) have been limited to small case series. Early reports suggest the Omicron (B.1.1.529) strain of SARS-CoV-2 (the dominant circulating US strain since the week of 12/25/2021) replicates more efficiently in the conducting airways. This may increase the risk of a croup phenotype in children as they have smaller airway calibers.

**Methods:** Description of the incidence, change over time, and characteristics of children with SARS-CoV-2 and upper airway infection (UAI) diagnoses within the National COVID Cohort Collaborative (N3C) before and during the rise of the Omicron variant. We compare the demographics, comorbidities, and clinical outcomes of hospitalized SARS-CoV-2 positive children with and without UAI.

**Results:** SARS-CoV-2 positive UAI cases increased to the highest number per month (N = 170) in December 2021 as the Omicron variant became dominant. Of 15,806 hospitalized children with SARS-CoV-2, 1.5% (234/15,806) had an UAI diagnosis. Those with UAI were more likely to be male, younger, white, have asthma and develop severe disease as compared to those without UAI.

**Conclusions:** Pediatric acute UAI cases have increased during the Omicron variant surge with many developing severe disease. Improved understanding of this emerging clinical phenotype could aid in therapeutic decision-making and healthcare resource planning.

## Introduction

SARS-CoV-2 can cause severe pediatric disease including acute COVID-19 and multisystem inflammatory syndrome in children (MIS-C)^1^. Published reports of SARS-CoV-2 causing laryngotracheobronchitis (commonly known as “croup”), however, have been limited to small case series^2^. Other coronaviruses (e.g. type NL63) are known to cause croup.

The Omicron (B.1.1.529) strain of SARS-CoV-2 became dominant in the U.S. during the week of 12/25/2021^3^. Early reports suggest Omicron may cause lower severity disease than the Delta variant^4^. This may be because Omicron replicates less efficiently in lung parenchyma and more efficiently in the conducting airways^5^. However, these mechanistic hypotheses have not been confirmed.

Young children are especially vulnerable to acute upper airway infection (UAI) because their airways are small and relatively collapsible. Inflammation from UAI can rapidly decrease air flow. Accordingly, croup is classically an early childhood disease. We conducted this retrospective cohort study to determine if acute UAI is more common as Omicron has become the dominant U.S. SARS-CoV-2 variant.

## Methods

We leveraged the National COVID Cohort Collaborative (N3C)^6^ and a pipeline we built for a NIH-funded pediatric COVID-19 dashboard (https://covid.cd2h.org/pediatrics-dashboard/) to conduct this study. Among all children in N3C <19-years-old with a positive SARS-CoV-2 test (polymerase chain reaction, antigen, or antibody), we identified those with a croup or tracheitis diagnosis. We included bacterial tracheitis because it can be difficult to distinguish from □— and can be a complication of □— viral croup. We compared groups using chi-square and Fisher exact tests for categorical variables and Mood’s Median test and t-tests for continuous variables. The N3C Data Enclave, data transfer from sites to N3C, and this analysis were approved under separate institutional review board protocols as documented elsewhere^1^.

## Results

SARS-CoV-2 positive UAI cases have increased as the Omicron variant has become dominant (Figure 1). During December 2021, we observed the largest monthly number thus far of hospitalized (N=32) and non-hospitalized (N=138) SARS-CoV-2 positive UAI cases. Of December 2021 hospitalizations, 2.5% (32/1302) had UAI. Overall, the 1/13/2022 N3C data release contains 15,806 hospitalized SARS-CoV-2 positive children, of whom 1.5% (234/15,806) had UAI (Table 1). Compared to those without UAI, those with UAI were more likely to be male (59.8% versus 50.4%, p=0.003), younger (2.4 versus 10.1 years, p<0.001), white (54.7% versus 43.3%, p=0.02), and have asthma (15% versus 10%, p=0.02). Children with UAI experienced severe disease (invasive ventilation, vasopressors, ECMO, or mortality) more often compared to children without UAI (31.6% versus 13.5%, p<0.001).

**Figure 1:**
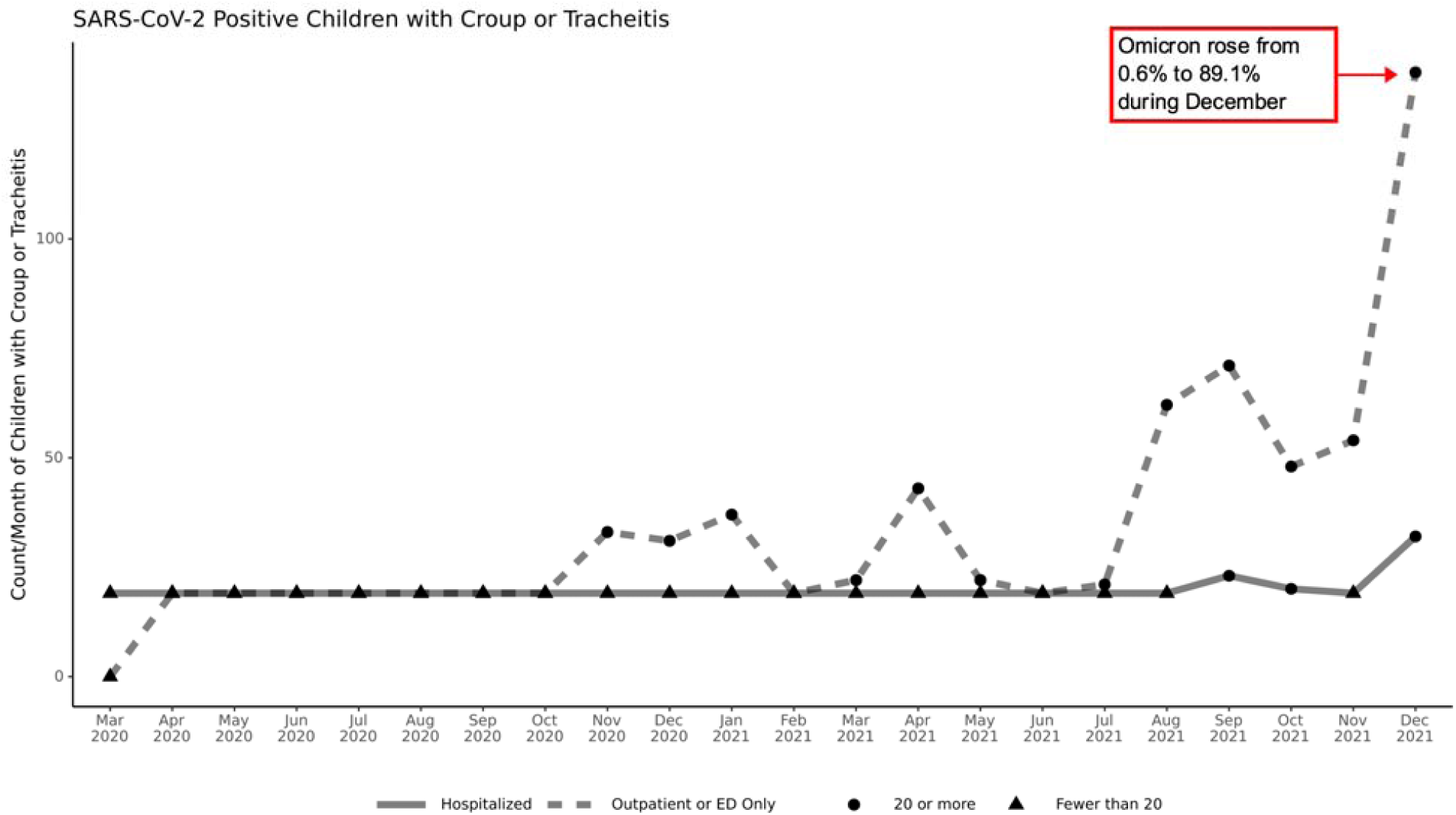
This figure shows the count per month of inpatient (solid line) and outpatient/Emergency Department (dotted line) encounters with a diagnosis of croup or tracheitis for children less than 19-years old with positive SARS-CoV-2 tests in the National COVID Cohort Collaborative (N3C) January 13th, 2022 data release. The percentage of SARS-CoV-2 samples found to be the Omicron strain is from the U.S. Centers for Disease Control COVID Data Tracker, available at https://covid.cdc.gov/covid-data-tracker/#variant-proportions and accessed January 19, 2022. Triangles represent individual counts 1-19 that are censored as per N3C policy and shown as n = 19 in the figure.

**Table 1:**
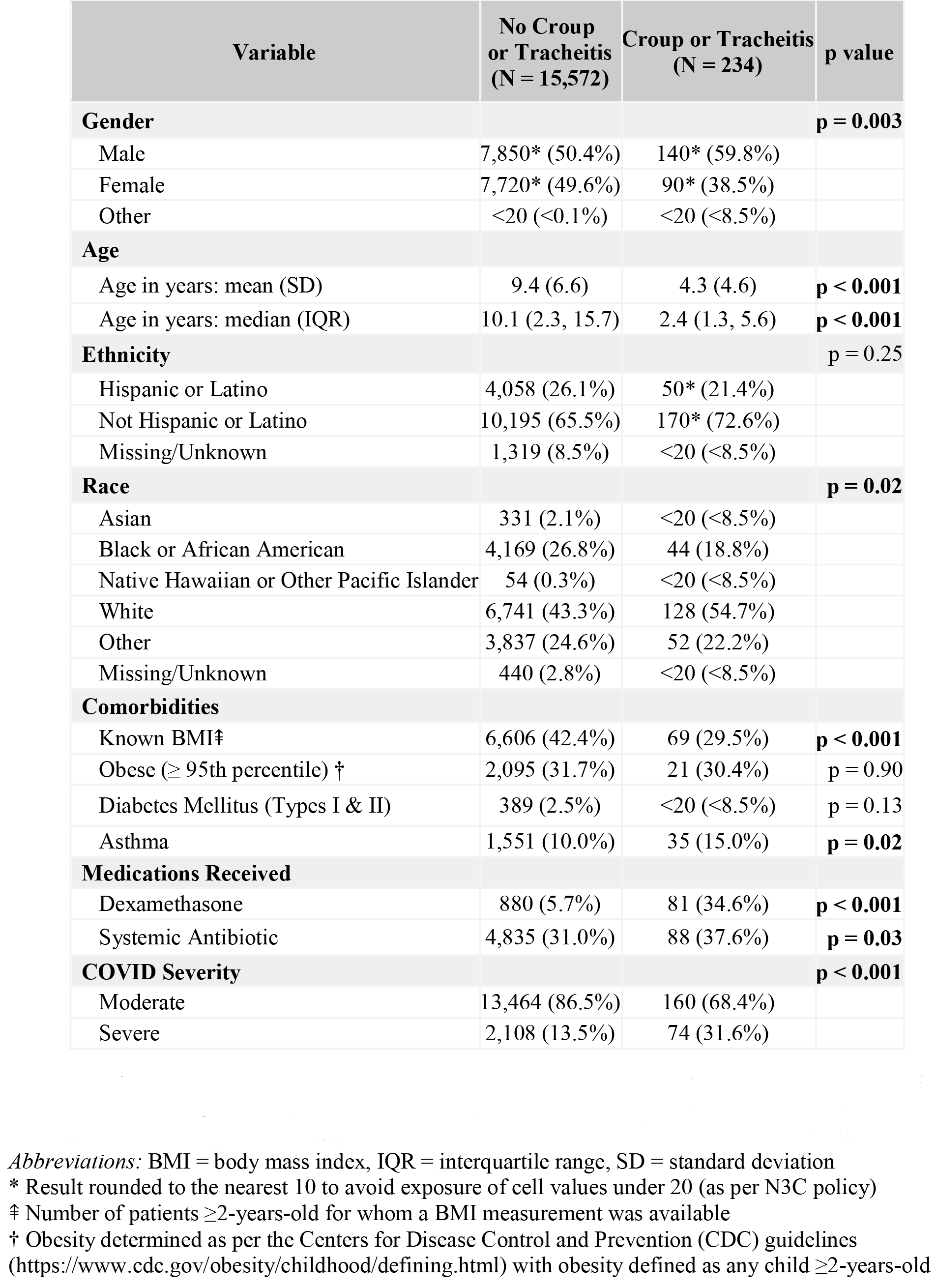
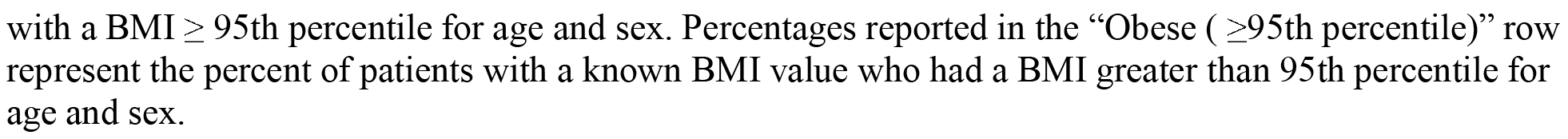
Children with COVID-19 with and without croup or tracheitis. This table shows demographic and clinical characteristics for children less than 19 years old with positive SARS-CoV-2 tests, with and without a diagnosis of croup or tracheitis. Per N3C policy, cells with less than 20 patients are reported as “<20 (x%)” where x is the percentage if n = 20.

| Variable | No Croup or Tracheitis (N = 15,572) | Croup or Tracheitis (N = 234) | p value |
| --- | --- | --- | --- |
| **Gender** |  |  | **p = 0.003** |
| Male | 7,850* (50.4%) | 140* (59.8%) |  |
| Female | 7,720* (49.6%) | 90* (38.5%) |  |
| Other | <20 (<0.1%) | <20 (<8.5%) |  |
| **Age** |  |  |  |
| Age in years: mean (SD) | 9.4 (6.6) | 4.3 (4.6) | **p < 0.001** |
| Age in years: median (IQR) | 10.1 (2.3, 15.7) | 2.4 (1.3, 5.6) | **p < 0.001** |
| **Ethnicity** |  |  | **p = 0.25** |
| Hispanic or Latino | 4,058 (26.1%) | 50* (21.4%) |  |
| Not Hispanic or Latino | 10,195 (65.5%) | 170* (72.6%) |  |
| Missing/Unknown | 1,319 (8.5%) | <20 (<8.5%) |  |
| **Race** |  |  | **p = 0.02** |
| Asian | 331 (2.1%) | <20 (<8.5%) |  |
| Black or African American | 4,169 (26.8%) | 44 (18.8%) |  |
| Native Hawaiian or Other Pacific Islander | 54 (0.3%) | <20 (<8.5%) |  |
| White | 6,741 (43.3%) | 128 (54.7%) |  |
| Other | 3,837 (24.6%) | 52 (22.2%) |  |
| Missing/Unknown | 440 (2.8%) | <20 (<8.5%) |  |
| **Comorbidities** |  |  |  |
| Known BMI‡ | 6,606 (42.4%) | 69 (29.5%) | **p < 0.001** |
| Obese (≥ 95th percentile) † | 2,095 (31.7%) | 21 (30.4%) | **p = 0.90** |
| Diabetes Mellitus (Types I & II) | 389 (2.5%) | <20 (<8.5%) | **p = 0.13** |
| Asthma | 1,551 (10.0%) | 35 (15.0%) | **p = 0.02** |
| **Medications Received** |  |  |  |
| Dexamethasone | 880 (5.7%) | 81 (34.6%) | **p < 0.001** |
| Systemic Antibiotic | 4,835 (31.0%) | 88 (37.6%) | **p = 0.03** |
| **COVID Severity** |  |  | **p < 0.001** |
| Moderate | 13,464 (86.5%) | 160 (68.4%) |  |
| Severe | 2,108 (13.5%) | 74 (31.6%) |  |

*Abbreviations:* BMI = body mass index, IQR = interquartile range, SD = standard deviation
\* Result rounded to the nearest 10 to avoid exposure of cell values under 20 (as per N3C policy)
‡ Number of patients ≥2-years-old for whom a BMI measurement was available
† Obesity determined as per the Centers for Disease Control and Prevention (CDC) guidelines (https://www.cdc.gov/obesity/childhood/defining.html) with obesity defined as any child ≥2-years-old
with a BMI ≥ 95th percentile for age and sex. Percentages reported in the “Obese ( ≥ 95th percentile)” row represent the percent of patients with a known BMI value who had a BMI greater than 95th percentile for age and sex.

## Discussion

Overall, pediatric acute UAI has increased during the Omicron variant surge. Nearly a third of affected children develop severe disease. This observed clinical phenotype of pediatric infection by the Omicron variant appears to confirm recent mechanistic reports.

A limitation of this analysis is that diagnosis codes will only be present for completed hospitalizations in N3C; children who are still hospitalized are not represented.

Although many children with acute UAI are managed as outpatients, those with severe croup and/or tracheitis are at risk of cardiac arrest from rapid-onset upper airway obstruction. They may require therapies typically provided in intensive care units including frequent administration of nebulized racemic epinephrine, helium/oxygen mixtures, and intubation. While SARS-CoV-2 pediatric UAI rates are not overwhelmingly high, understanding this new clinical phenotype is important to health systems under severe strain. Anticipation of the potential for acute upper airway obstruction may guide therapeutic decision-making and hospital planning for available equipment and personnel.

## Data Availability

All data and analyses utilized in the present study are available within the N3C data enclave and accessible following N3C registration and submission of a Data Use Request. For more information visit:
https://ncats.nih.gov/n3c/about/applying-for-access

## Acknowledgements

The N3C was funded by NCATS grant number U24 TR002306 and other support as documented at https://covid.cd2h.org/acknowledgements. This analysis was supported by NICHD grant number R01HD105939-01S1.

## References

1. Martin B, DeWitt PE, Russell S, et al. (in press). Children with SARS-CoV-2 in the National COVID Cohort Collaborative (N3C). JAMA Netw Open. 2022.

2. Venn AMR, Schmidt JM, Mullan PC. Pediatric croup with COVID-19. Am J Emerg Med. 2021;43:287 e281–287 e283.

3. Centers for Disease Control and Prevention. COVID Data Tracker: Variant Proportions. https://covid.cdc.gov/covid-data-tracker/#variant-proportions. Published 2022. Accessed Jan 18, 2022.

4. Ulloa AC, Buchan SA, Daneman N, Brown KA. Early estimates of SARS-CoV-2 Omicron variant severity based on a matched cohort study, Ontario, Canada. medRxiv. 2021.

5. Chan MC, Hui KP, Ho J, et al. SARS-CoV-2 Omicron variant replication in human respiratory tract ex vivo. Dec 2021, PREPRINT (Version 1) available at Research Square https://doi.org/10.21203/rs.3.rs-1189219/v1

6. Haendel MA, Chute CG, Bennett TD, et al. The National COVID Cohort Collaborative (N3C): rationale, design, infrastructure, and deployment. Journal of the American Medical Informatics Association. 2021;28(3):427–443.

